# Standardization of a flow cytometry SARS-CoV-2 serologic test

**DOI:** 10.1101/2021.01.29.21250770

**Authors:** Carl Simard, Jonathan Richard, Renée Bazin, Andrés Finzi, Patrick Trépanier

## Abstract

The SARS-CoV-2 virus is the causing agent of the coronavirus disease 2019 (COVID-19) pandemic which is responsible for millions of deaths worldwide. The development of the humoral response to the virus has been the subject of intensive research and development. A flow cytometry-based assay using native full-length SARS-CoV-2 Spike protein expressed in 293T cells was recently proposed as a complementary seropositivity determination assay.

The aim of our study was to further develop the flow cytometry assay for potential use as a confirmatory test and to standardize its parameters and results for reliable inter-laboratory use. We have optimized the protocol, established the Receiving Operating Characteristic (ROC) curve and tested reproducibility using pre-COVID plasma samples and convalescent, SARS-CoV-2 individual plasma samples.

The flow-based assay was simplified and standardized by cultivating the 293T cells in suspension and expressing results in Mean Equivalent Soluble Fluorochrome (MESF) using an internal antibody positive control. The ROC curve was determined with an area under the curve (AUC) of 0.996 and the assay specificity and sensitivity were established at 100% and 97.7% respectively. Reproducibility was good as determined on multiple cytometers, on different days, and with data acquisition as far as 72h post-staining. The optimized and standardized assay could be used as a high throughput confirmation confirmatory assay in flow cytometry laboratories involved in serological testing.

## Introduction

The coronavirus disease 2019 (COVID-19) caused by the severe acute respiratory syndrome coronavirus 2 (SARS-CoV-2) pandemic has caused over 2.1 million deaths worldwide as of January 2021 (https://www.worldometers.info/coronavirus). It has been shown that the virus Spike (S) glycoprotein binds the human angiotensin-converting enzyme 2 (ACE2) through its receptor-binding domain (RBD)^1,2^. Intensive work has been done since the beginning of the pandemic to develop reliable, sensitive and specific immunosorbent assays to characterize the humoral response of humans to the virus. The seropositivity determination allows for seroprevalence studies^3^, for the characterization of plasma in the development of convalescent plasma therapy^4^ and could be used for the study of the overall antibody responses over time^5^, which include vaccine response persistence in due time.

Recently, Anand et al. suggested an innovative flow cytometry approach using the native full-length SARS-CoV-2 S protein stably expressed by 293T cells (Anand et al., Transfusion, 2021, in press)^6^. This approach circumvents the potential antibody detection limitations consequential of using RBD peptides for seropositivity determination. This protocol also addresses the potential supply chain plastic shortage some country may face by allowing testing using variable tube formats.

Given that the described flow cytometry assay may be of use in many laboratories worldwide, we herein propose a simplified and standardized protocol destined to groups interested in importing the assay within their flow cytometry operations. This paper presents a standardized protocol in order to import the semi-quantitative assay in a lab, a proposed Mean Equivalent Soluble Fluorochrome (MESF)-to-control-antibody ratio approach to interpret the data and the positivity threshold determination and reproducibility using convalescent donor samples. Overall, this adaptation should facilitate the integration of such a technique in any flow laboratory and sets ground for further development.

## Material and methods

### Plasma samples

Pre-COVID plasmas were obtained during regular blood donations in the province of Quebec, Canada, before December 20^th^ 2019. It is assumed that the disease was absent from this area before this date. Convalescents plasmas were randomly chosen from adult participants in the CONCOR-1 study (#NCT04348656) having received an official diagnosis of COVID-19 by the Québec Provincial Health Authority confirmed by polymerase chain reaction (PCR). All patients have been symptomatic without hospitalisation and free of symptoms for at least two weeks before donation. All donors gave consent to participate in this research project which was approved by the Héma-Québec Research Ethic Committee.

### Spike-expressing 293T cells

The 293T cells stably expressing the SARS-CoV-2 Spike protein (293T.SARS-CoV-2 Spike) were previously described (Anand et al., Transfusion, 2021, in press)^6^. The cells, as well as non-transformed 293T control cells, were adapted to suspension culture in 293 SFM II medium (ThermoFisher #11686029) as per manufacturer protocol.

### Staining and flow cytometry analysis

A detailed protocol is provided as supplementary material. Briefly, a mix of 293T-SARS-CoV-2 Spike stable cell line and 293T control cells were stained with an anti-RBD CR3022 monoclonal Ab (50 ng/ml) (graciously provided by A. Finzi) as control or plasma (1:400 dilution) for 20 minutes at room temperature. Optimal plasma dilution was determined by titration curve (Supplemental Data). After washing, AlexaFluor-647-conjugated goat anti-human IgG+IgA+IgM Abs (Jackson Immunoresearch 109-605-064) diluted 1:400 (final concentration of 3.75 µg/ml) was used as detection antibody for 20 minutes at room temperature. Optimal antibody concentration was determined by titration curve (Supplemental Data). After washing, samples were fixed for 10 minutes in formaldehyde 2% in DPBS before being centrifuged and resuspended in DPBS-BSA 0.2% (BD 554657). Data acquisition was done on Accuri C6 flow cytometers in standard configuration with compensations obtained from calibration beads. Results were transformed in Alexa Fluor 647 MESF using a standard curve from beads stained with a known quantity of fluorochrome (Quantum MESF Alexa Fluor 647, Bang Laboratories, #647) (Supplemental Data). Results were reported as the ratio of the sample MESF on the CR3022 control MESF.

### Statistical analyses

Statistical analyses were done using R 4.0.3, using package OptimalCutpoints 1.1-4^7^. The determination of the optimal seropositivity threshold was based on the Youden’s index.

## Results

The distribution of results expressed as MESF to CR3022 control antibody ratio from the plasma of N=56 pre-COVID donors and of N=44 convalescent donors is shown in Figure 1A. The data obtained from the assay show a clear separation of anti-S negative and positive groups. The means of MESF ratio were 0.30 ± 0.14 and 13.95 ± 14.39 for pre-COVID and convalescent samples respectively. In order to assess the performance of the test as a classifier for COVID-19 antibodies presence and determine a positivity threshold, we established a Receiving Operating Characteristic (ROC) curve (Figure 1B). The area under the curve (AUC) of 0.996 indicates excellent discrimination between negative and positive samples. The optimal threshold point of 1.1 determined from Younden’s index has a sensitivity of 97.7% (IC95: 88,0% - 99,9%) and a specificity of 100% (IC95: 93,6% - 100%). Therefore, a MESF ratio greater or equal to 1.1 relative to CR3022 50ng/ml is considered to be indicative of seropositivity (Figure 1A, dashed grey line).

**Figure 1:**
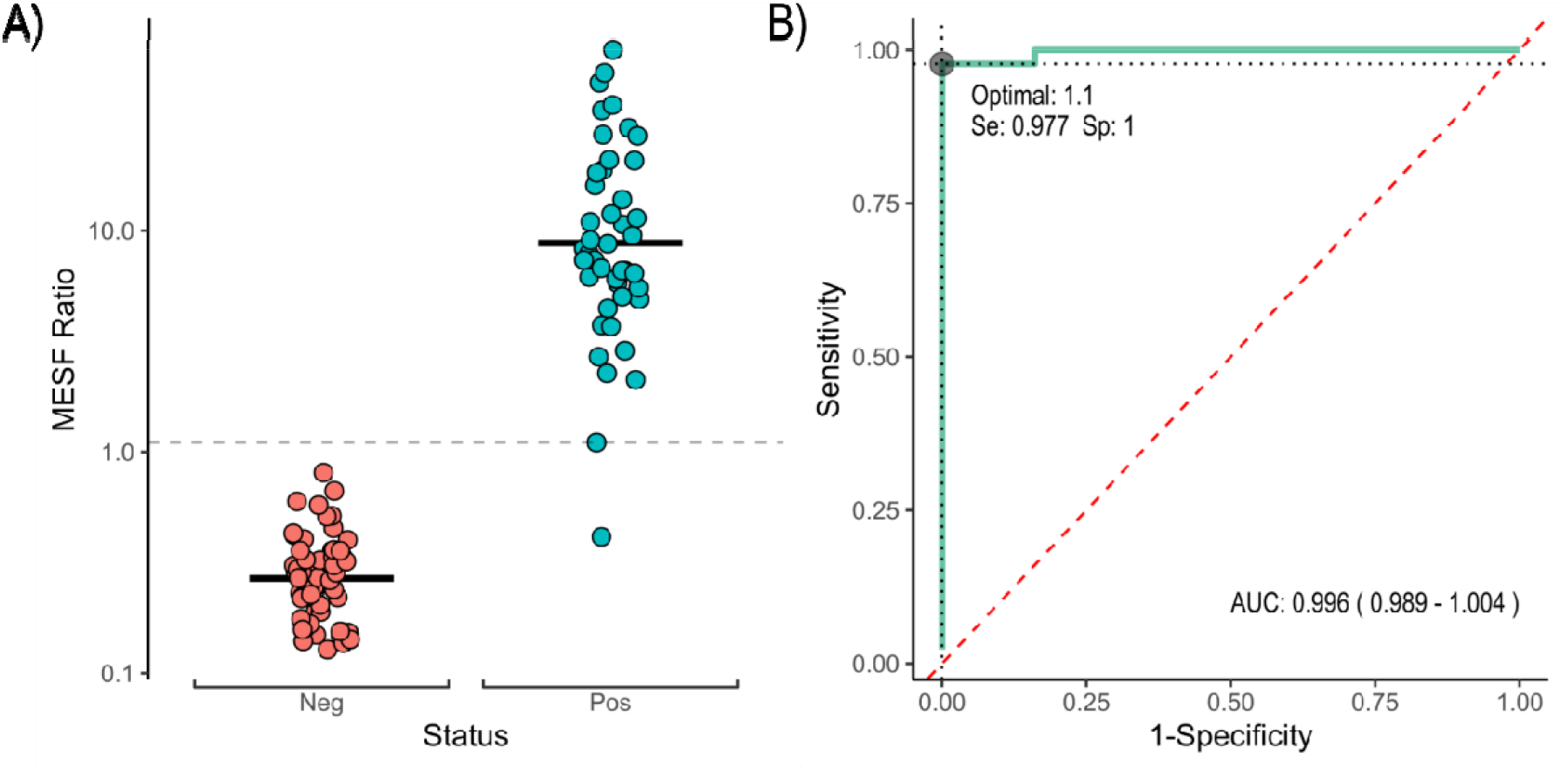
(A) Distribution of MESF ratios relative to CR3022 (50 ng/ml) control for pre-COVID (Neg, red dots), n=56, and convalescent plasmas (Pos, green dots), n=44. The dotted line represents the positivity threshold. Black bars are the means for each group (0.30 and 13.95 for negative and positive groups respectively). (B) ROC curve (green) and optimal threshold point determined by Youden’s index. The red dashed line is the line of no discrimination.

In order to validate the standardized protocol, the reproducibility was evaluated by analyzing samples on three different Accuri C6 flow cytometers, on two different days (Figure 2A). The mean CV of 0.070 supports highly reproducible results. In order to accommodate higher volume of testing and more flexibility, the post-staining stability was evaluated by analyzing samples stored at 4°C in the dark after 24h and 72h (Figure 2B). The mean CV of 0.023 suggests that samples can be analyzed up to 72h after staining without any loss of signal, and therefore, allows for an extended sample preparation period before acquisition.

**Figure 2:**
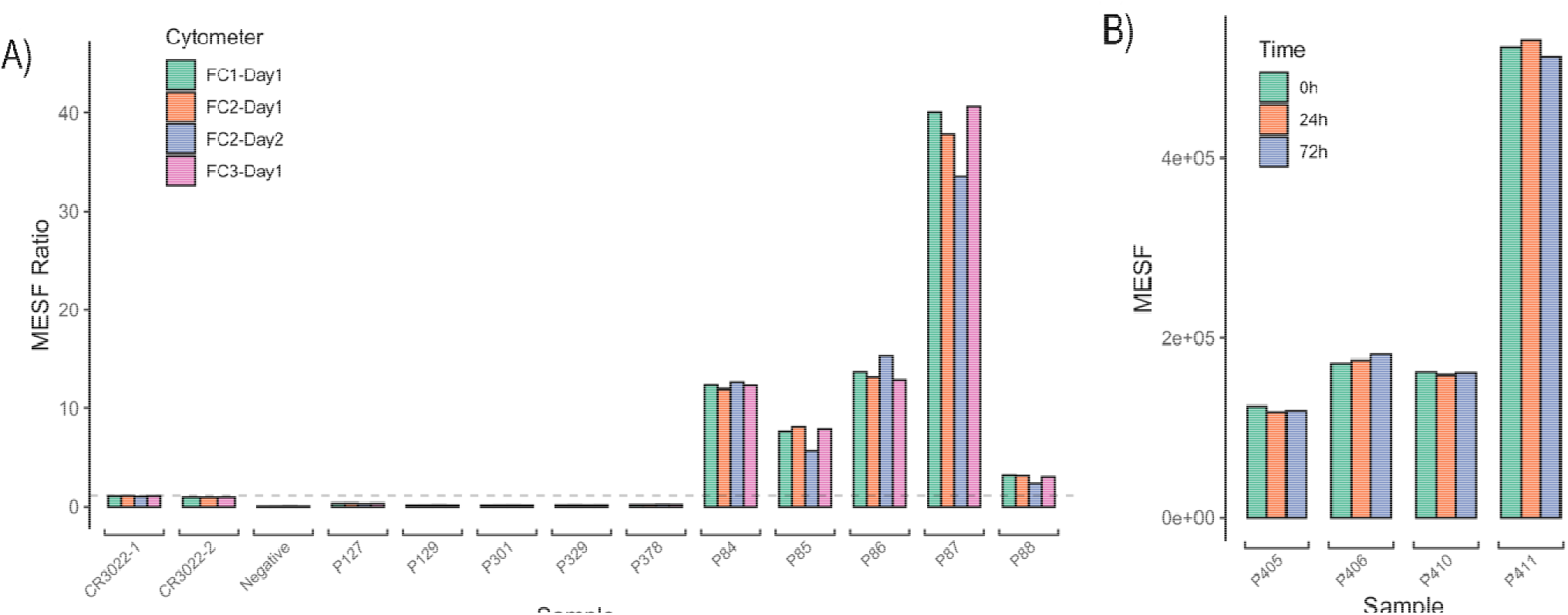
Reproducibility of results for samples analyzed on (A) three different flow cytometers (identified as FC1, FC2 and FC3) on two different days (Day1 and Day2). Negative are samples without plasma, P127 to P378 are pre-COVID plasmas, P84 to P88 are convalescents plasmas. The dashed line represents the previously determined threshold value of 1.1 MESF Ratio. (B) MESF of four convalescents plasmas analyzed immediately (0h), 24h and 72h after staining.

## Discussion

The SARS-CoV-2 pandemic has accelerated the development of many serodiagnostic platforms and inspired the establishment of alternative methods of analysis to circumvent some technical and/or supply chain challenges, such as the flow cytometry-based assay to detect SARS-CoV-2 Spike specific antibodies described by Anand et al. (Transfusion, 2021, in press)^6^. In this study, we report a protocol optimization regarding the cell culture in suspension and the quantitative measurements using a ratio of MESF and of an internal control, and propose standardization steps to facilitate the implementation of the method in any flow cytometry laboratory. In addition, we present the positivity threshold determination by comparing non-COVID-19 to confirmed-COVID-19 samples and describe how sensitive (97.7%) and specific (100%) the flow cytometry assay is. Finally, with the suggested standardization steps, we support the potential use of this flow assay as a quick and potentially high throughput confirmation test in a non-clinical setting.

Given our understanding of the challenges associated with flow cytometry and our experience in inter-laboratory standardization of flow assays, we ought to provide the community with a standardized baseline which could be adapted for further characterization of the SARS-CoV-2 humoral response. For example, laboratories who only have residual or low sample volume could adapt this assay to measure the IgG1/2/3/4, IgA and IgM distinct response all within one tube using multiple fluorescent antibodies. Also, while using 293T cells in suspension facilitated cell culture and time management, our adaptation only allows for about 20 samples to be done from start to finish, by one technician, during a regular day. This capacity suited our testing requirement as we mostly used the technique to resolve unclear, near threshold ELISA results. Laboratories who require a higher throughput could setup an automated sampler protocol on their flow cytometer, and/or use our described 72 hours post-staining stability to batch flow cytometry data acquisition. Overall, our study demonstrates that the adapted flow cytometry assay is robust enough to be used as a seropositivity confirmation assay. Altogether, our findings set the standardization basis for a SARS-CoV-2 flow cytometry assay using Spike-expressing transfected 293T cells and confirm the possibility of using the assay as an indicator of the serological response to the virus.

## Data Availability

No additionnal datasets are available.

## Acknowledgements

The authors are grateful to the convalescent plasma donors who participated in this study and the Héma-Québec team involved in convalescent donor recruitment and plasma collection. We thank Dr. M. Gordon Joyce (U.S. MHRP) for the monoclonal antibody CR3022.

## Fundings

This work was supported by le Ministère de l’Économie et de l’Innovation du Québec, Programme de soutien aux organismes de recherche et d’innovation and by the Fondation du CHUM to A.F. This work was also supported by Canada’s COVID-19 Immunity Task Force (CITF), in collaboration with the Canadian Institutes of Health Research (CIHR) and a CIHR foundation grant #352417 to A.F. A.F. is the recipient of Canada Research Chair on Retroviral Entry no. RCHS0235 950-232424. The funders had no role in study design, data collection and analysis, decision to publish, or preparation of the manuscript.

### Abbreviations

ACE2: angiotensin-converting enzyme 2
AUC: area under the curve
COVID-19: coronavirus disease 2019
MESF: Mean Equivalent Soluble Fluorochrome
PCR: polymerase chain reaction
ROC: receiving operating characteristic
RBD: receptor-binding domain

## Notes

### Competing Interest Statement

The authors have declared no competing interest.

### Funding Statement

Fundings
This work was supported by le Ministere de l Economie et de l Innovation du Quebec Programme de soutien aux organismes de recherche et d innovation and by the Fondation du CHUM to A.F. This work was also supported by Canada s COVID-19 Immunity Task Force (CITF), in collaboration with the Canadian Institutes of Health Research (CIHR) and a CIHR foundation grant #352417 to A.F. A.F. is the recipient of Canada Research Chair on Retroviral Entry no. RCHS0235 950-232424. The funders had no role in study design data collection and analysis, decision to publish or preparation of the manuscript.

### Author Declarations

All donors gave consent to participate in this research project which was approved by the Hema-Quebec Research Ethic Committee.

