## Supplemental Data for "Standardization of a flow cytometry SARS-CoV-2 serologic test"

### **Supplemental Material**

#### Detailed protocol

The principles behind this method are explained extensively in Anand at al. Briefly, the plasma to be tested is put in presence of cultured cells expressing the SARS-COV2 Spike protein in native form (293T-SARS-CoV-2 cell line). Antibodies presents in the plasma, if present, will bind this protein at the surface of the cells. Their presence could then be detected by using a fluorescent antibody against human IgM-IgG-IgA. The non-transformed 239T cells are used as control to identify GFP-positive 293T-SARS-CoV-2 Spike cells.

Count 293T and 293T-SARS-CoV-2 Spike cell cultures.

Mix 50 000 cells of each culture per sample to be analyzed (total of 100 00 cells / sample).

Centrifuge at 200g for 5 minutes, room temperature.

Discard supernatant and resuspend the cell pellet in PBS-BSA 0.2% in a total volume of 100 µl per sample to be analyzed.

For each sample, add 1 µl of plasma diluted 1 :4 in PBS-BSA 0.2%..

Add 1 µl of CR3022 5 µg/ml to the duplicate control tubes.

Add 99 µl of cell suspension to each sample and control tubes and mix.

Incubate for 20 minutes at room temperature protected from light.

Centrifuge at 200g for 5 minutes, room temperature.

Discard supernatant and wash with 500µl of cold PBS-BSA 0.2%

Centrifuge at 200g for 5 minutes, room temperature.

Add 100 µl of Alexa Fluor 647 goat anti-human IgA+IgG+IgM (H+L) (Jackson Immunoresearch 109-605-064) antibody diluted 1:400 in PBS-BSA 0.2% The final concentration is 3.75 µg/ml.

Incubate for 20 minutes at room temperature protected from light.

Centrifuge at 200g for 5 minutes, room temperature.

Discard supernatant and wash with 500 µl of cold PBS-BSA 0.2%.

Centrifuge at 200g for 5 minutes, room temperature.

Discard supernatant and resuspend in DPBS-formaldehyde 2%.

Incubate for 10 minutes at room temperature protected from light.

Centrifuge at 200g for 5 minutes, room temperature.

Discard supernatant and resuspend in PBS-BSA 0.2%.

If needed, at this stage the samples can be kept for at least 72h at 4°C, protected from light before proceeding to flow cytometry analysis.

Analyze on flow cytometer and measure the Alexa Fluor 647 median fluorescence intensity (MFI) of the GFP positive cell population. The MFI is converted to MESF using the calibration curve from Quantum MESF Alexa Fluor 647 beads as per manufacturer instructions and using the calibration template provided with the kit. For each sample, the MESF is divided on the mean MESF of control tubes to get the ratio.

A typical gating from a convalescent plasma is shown below:

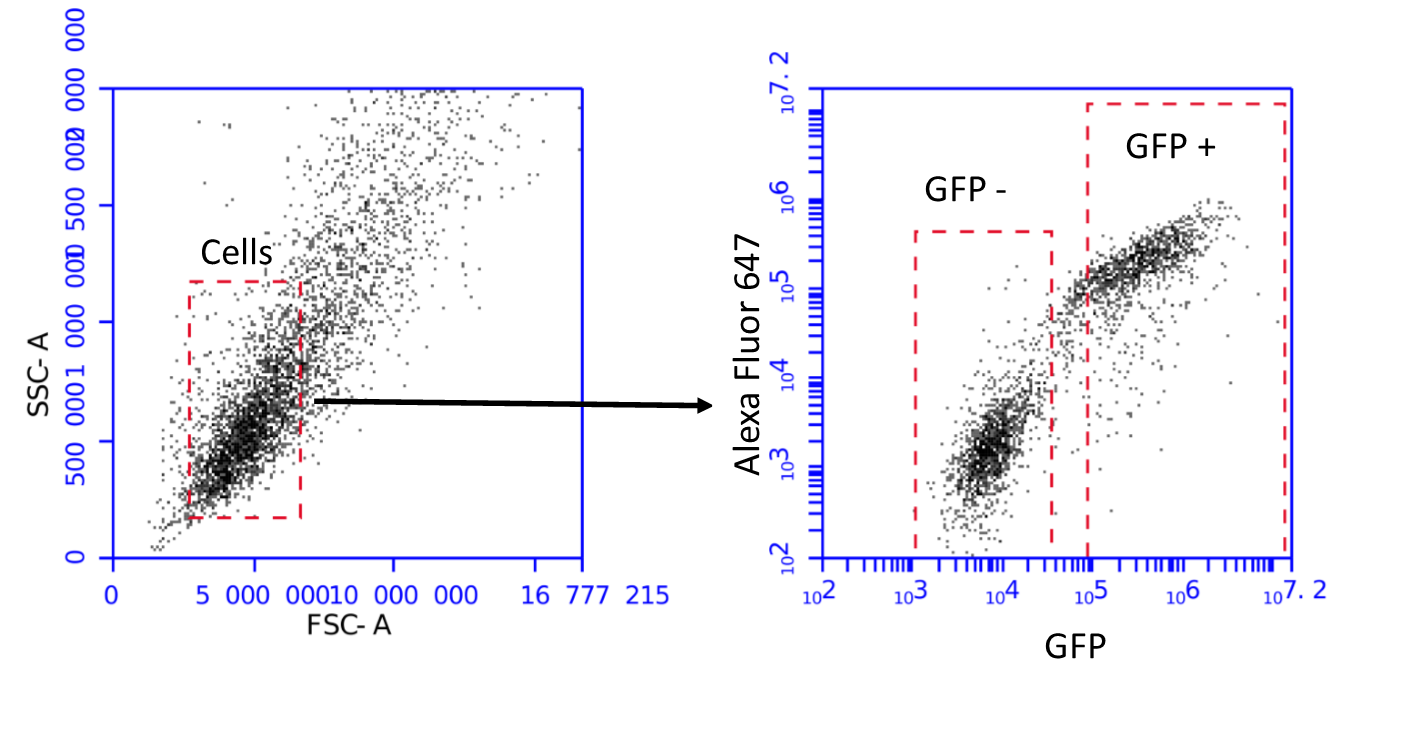

The cell population is fist delimited on a FSC – SSC plot. The selected cells are then reported on a Alexa Fluor 647 – GFP plot. The median fluorescence value of the GFP positive population is determined and converted to MESF as described above. The GFP – population is used to confirm the SARS-COV-2 cell line is still expressing the spike protein, which is coexpressed with GFP.

#### CR3022 Titration

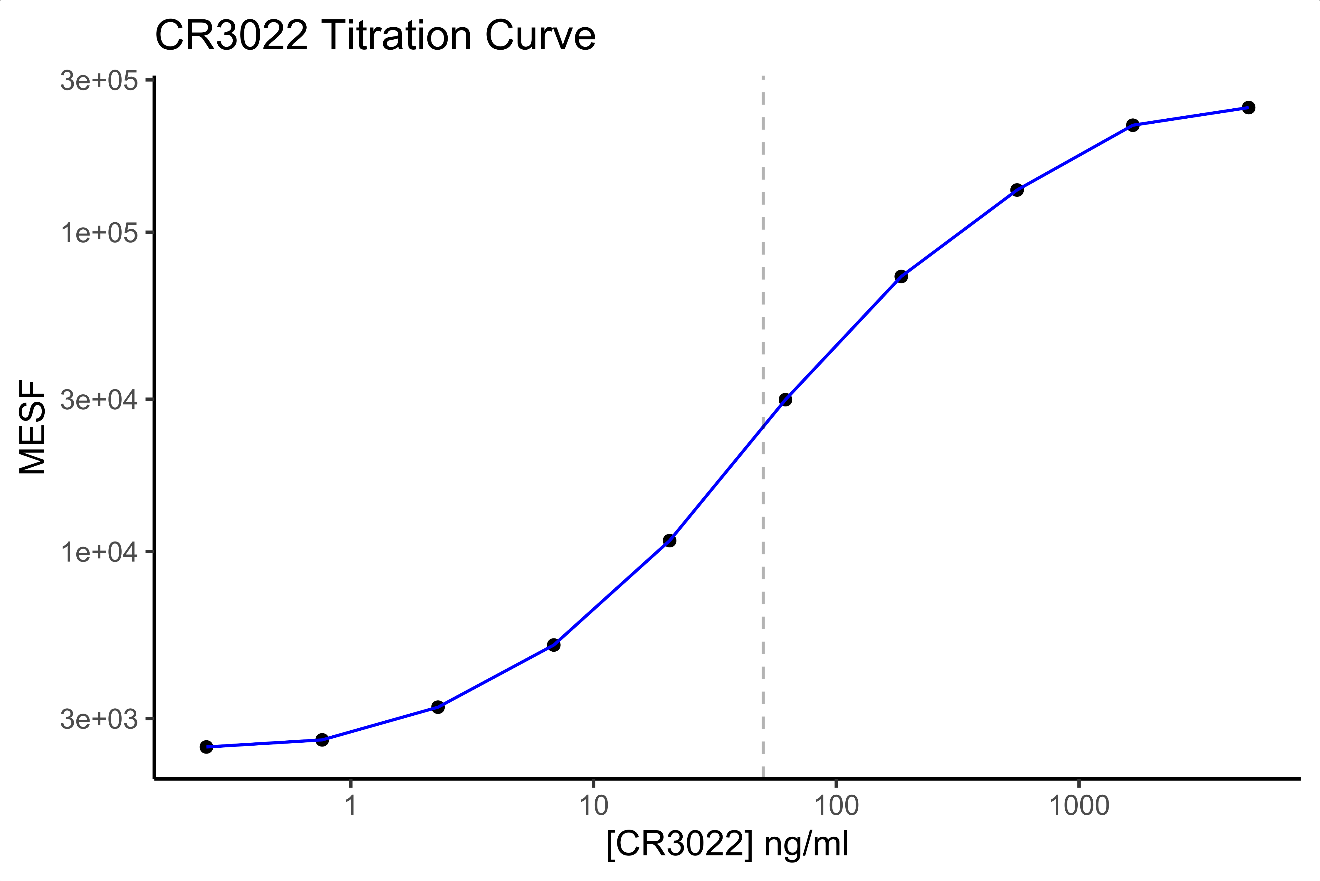

Titration of CR3022 was performed by 1:3 serial dilution of CR3022 antibody in PBS-BSA 0.2%. The dotted line corresponds to the concentration of 50 ng/ml used as an internal calibration control. This concentration being in the middle of the linear range (from about 5 ng/ml to 500 ng/ml) it should allow, as a calibrator, the optimal compensation for inter-assay variation.

#### Antibody titration

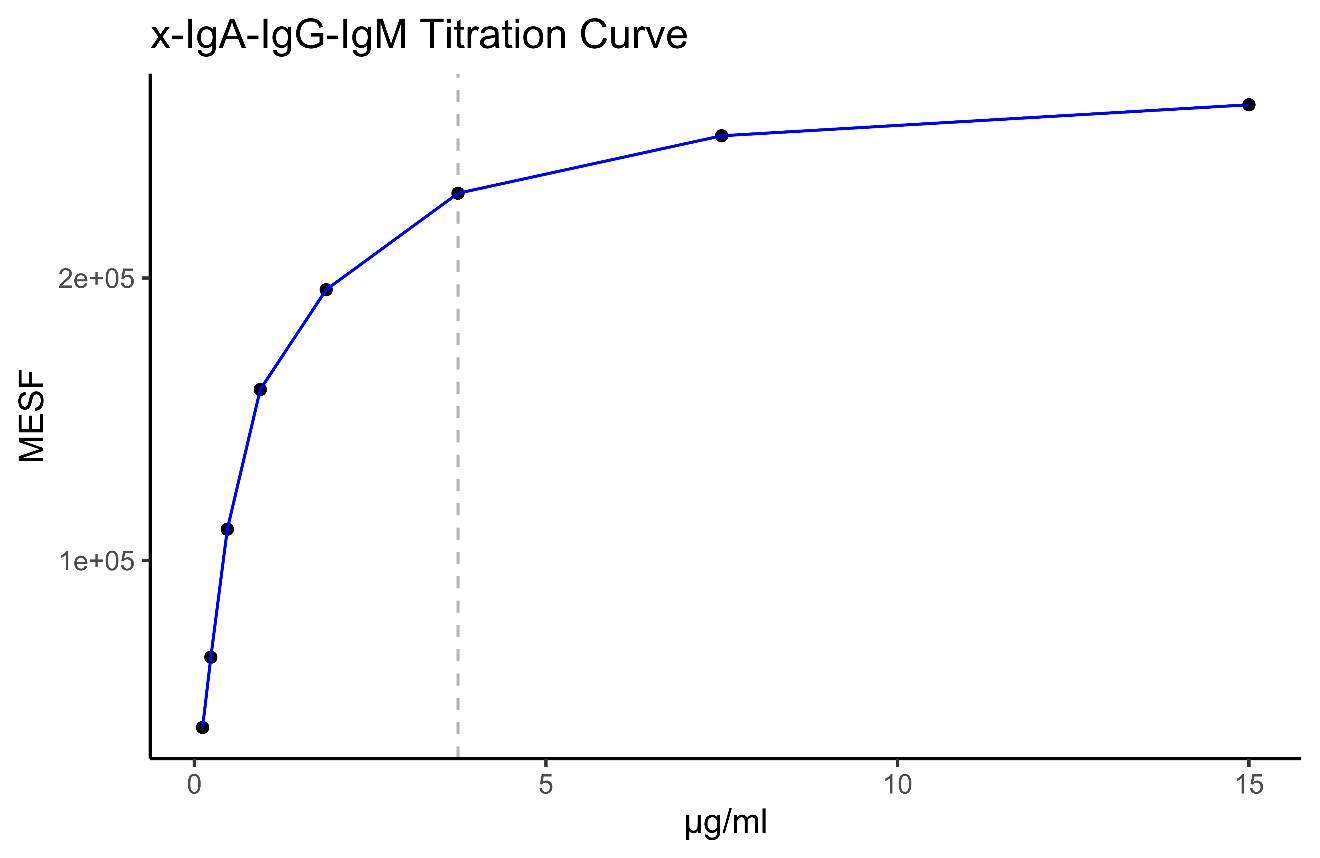

The Alexa Fluor 647 goat anti-human IgA+IgG+IgM (H+L) (Jackson Immunoresearch 109-605-064) antibody was titrated by serial dilution against 293T-CoV2 cells and CR3022 at the saturating concentration of 500 ng/ml. The dashed grey line corresponds to a dilution of 1 :400 (3.75 µg/ml).

#### Plasma titration

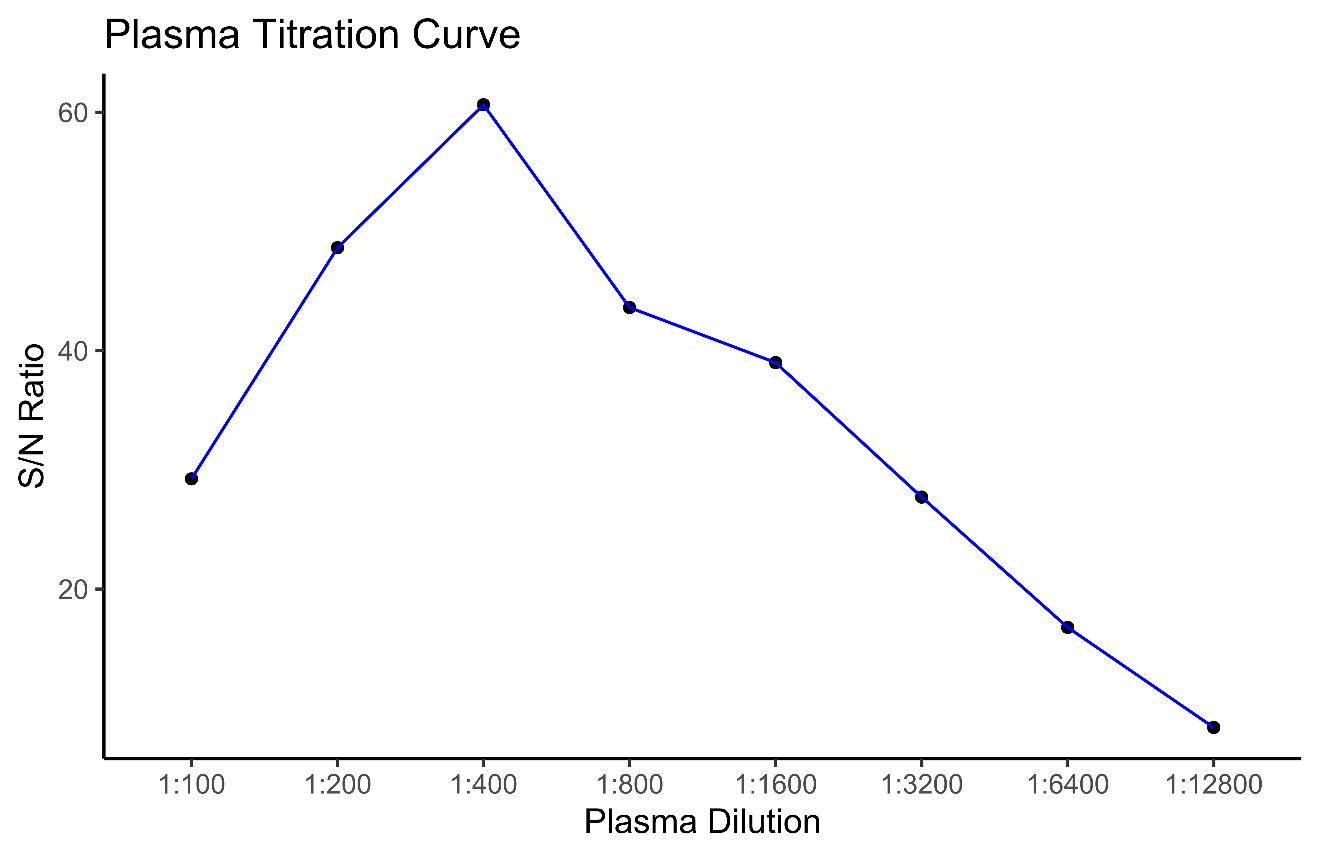

The plasma dilution was optimized based on a titration curve of signal noise ratio against plasma dilution. The signal to noise ratio was determined by dividing the MESF value of a convalescent plasma by the MESF value of a negative control consisting of a commercial pool of plasma (Seracon) at the same dilution.
